# Optimized sampling method for fecal microbiome and metabolome preservation under room temperature

**DOI:** 10.1101/2023.05.08.23289643

**Authors:** Tatsuhiro Nomaguchi, Yohsuke Yamauchi, Yuichiro Nishimoto, Yuka Togashi, Masaki Ito, Felix Salim, Kota Fujisawa, Shinnosuke Murakami, Takuji Yamada, Shinji Fukuda

## Abstract

**Background:** The relationship among the human gut microbiome, microbially produced metabolites, and health outcomes remains of great interest. To decrease participant burden, room-temperature storage methods for fecal samples have become increasingly important. However, kits for storing the fecal microbiome and metabolome have not been well explored. We hypothesized that storing fecal samples by drying them with silica gel may be suitable.

**Objectives:** The objective was to evaluate the performance of storage at room temperature by drying feces for subsequent examination of the microbiome, microbial pathways, and the metabolome.

**Methods:** Feces from ten healthy adults (6 male and 4 female) were sampled and immediately processed, as controls, and stored at room temperature in an incubator, on an FTA card, in RNAlater, or dried by silica gel. Storage at room temperature continued for 7 days. Drying by the silica gel method was assessed for 14 days. The fecal microbiome was assessed by sequencing the bacterial 16S ribosomal RNA-encoding gene (V1-V2 region), fecal microbial pathway profiles were analyzed by whole-genome shotgun metagenomics, and fecal metabolome profiles were analyzed using capillary electrophoresis time-of-flight mass spectrometry (CE-TOFMS).

**Results:** Qualitative and β-diversity analyses of the microbiome, microbial pathways, and the metabolome showed that drying by silica gel were closest to those immediately after processing. When focusing on the abundances of individual microbes, microbial pathways, and metabolites, some were found to be significantly different. However, the intra-method ranking of individual items showed that 100%, 87-97%, and 63-76% of microbes, microbial pathways, and metabolites, respectively, were significantly correlated between silica gel preserving and immediately processing method.

**Conclusions:** The results showed that fecal sample drying could be effectively used for the preservation of the fecal microbiome and metabolome.

## 1 Introduction

The gut microbiome and microbiome-derived metabolites are strongly associated with host health. Some examples include not only intestinal diseases such as inflammatory bowel diseases [1], *Clostridioides difficile* infection [2], and colorectal cancer [3] but also systemic diseases such as glucose intolerance [4], and atherosclerosis [5]. Studies have revealed the importance of evaluating the gut environment and thus the development of evaluation methodology, including collection methods.

In the academic field, the standard fecal sample collection method for evaluating the gut microbiome and metabolome is freezing specimens immediately after collection because the microbial composition in feces can change considerably within a short period at room temperature [6]. However, immediate freezing requires freezer or laboratory equipment, which is inconvenient for routine clinical practice and for obtaining samples in fieldwork locations such as jungles. Therefore, methods to preserve fecal samples without freezing have been explored. In previous studies, commercially available sampling kits and/or reagents that were considered capable of storing the fecal microbiome at room temperature were evaluated [7–9]. While these studies have suggested the potential of some kits for preserving the microbiome composition, there is currently no methodology for preserving both the microbiome and metabolome. Furthermore, current sampling kits frequently contain fixing reagents with high salt contents that are not safe to use for collection outside the laboratory, also causing difficulty for metabolomic measurement by mass spectrometry.

For alternate preservation methods of the microbiome and metabolome composition in fecal samples, we focused on drying. Drying, or more precisely, low water activity is a classic but reliable method to inhibit microbial proliferation and has been used as a food storage technique for centuries [10]. Although some microbes, such as spore-forming bacteria, are resistant to low water activity and are able to persist, low water activity is able to prevent microbial growth [11]. In addition, sample drying with desiccant has been used as a common DNA sampling method by field biologists, and several studies have reported its DNA preservation potency [12,13]. Therefore, immediate drying with desiccant may have great potential for the preservation of microbial community composition and microbial DNA.

On the other hand, there is a concern that the drying process may cause loss of some volatile metabolites in fecal samples. Gut microbiome constituents metabolize dietary fibers reaching the large intestine to produce a variety of organic acids, including short-chain fatty acids (SCFAs), which have been reported to be important factors for many health and disease conditions [14,15]. These organic acids include some volatile substances that would be lost while the fecal samples dry. However, a previous study reported that fecal SCFA concentrations are stable after the freeze-drying procedure, which may suggest the possibility that fecal metabolites may also be preserved in feces during desiccant drying [16].

In this study, we aimed to evaluate a stool preservation method based on drying with silica gel, a globally common desiccant, as a method of fecal sample preservation at room temperature that can maintain the microbiome and metabolome profiles. As a result, qualitative and β-diversity analyses of the microbiome, microbial pathways, and metabolome showed that drying by silica gel was the closest to immediate freezing compared to other preservation methods, such as RNAlater and FTA cards. In addition, the intramethod ranking of individual items showed that 100%, 87-97%, and 63-76% of microbes, microbial pathways, and metabolites, respectively, were significantly correlated with the immediate freezing method. These results showed that drying fecal samples with silica gel has great potential for the preservation of the fecal microbiome and metabolome.

## 2 Methods

### 2.1 Trial design and sample collection

Storage of human fecal samples was tested using several methods (Fig. 1).

**Fig. 1.**
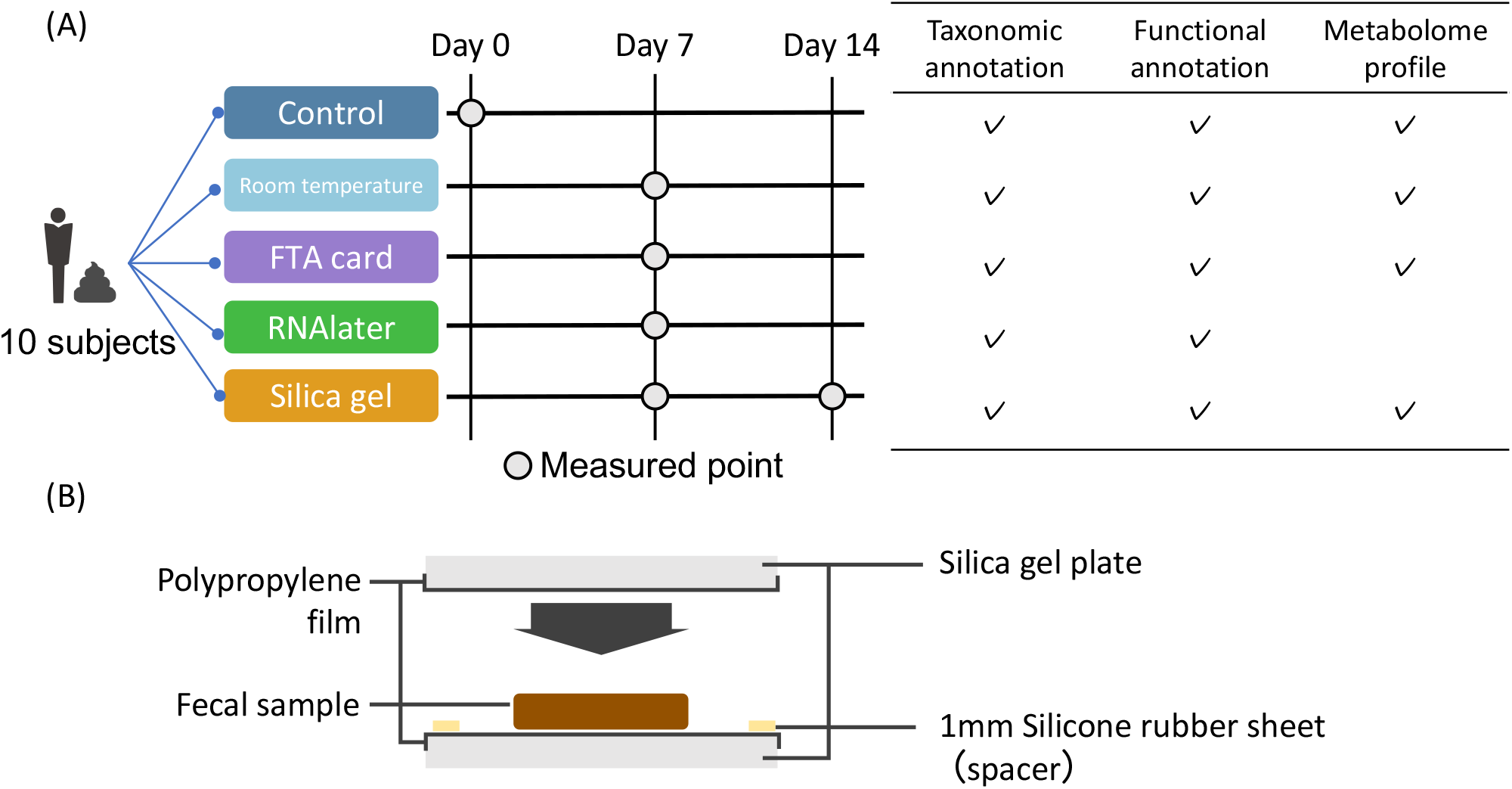
Overview of the study. (A) Overview of the study. (B) Schematic diagram showing how the fecal sample was stored with silica gel.

- Immediately frozen with liquid nitrogen after fecal sample collection and then used for DNA and metabolite extraction. Processed as a positive control in this study (abbreviated method name: Control).
- Stored at 25°C (room temperature maintained by incubator) as a negative control (abbreviated method name: Room temp., stored at room temperature).
- Stored at 25°C with an FTA™ card (GE healthcare, USA) (abbreviated method name: FTA card). The application process was as follows: 1) the fecal sample was applied onto the FTA card, 2) the sample was allowed to dry for one hour at room temperature (official protocol), and 3) the FTA card was placed into a hermetically sealed bag and stored in the incubator.
- Stored at 25°C with RNAlater (Qiagen, Netherlands) (abbreviated method name: RNAlater).
- Stored at 25°C with MGKit (Metagen, Japan) (abbreviated method name: Silica gel). The application process was as follows: 1) a fecal sample was sandwiched with a predried silica gel plate (30 × 40 × 3 mm^3^)that was wrapped with microporous polypropylene film, allowing dried feces to be easily removed, and a 1 mm thick-silicone rubber sheet was also sandwiched to ensure that the fecal sample was stored at a certain thickness to standardize the drying efficiency; and 2) silica-sandwiched feces was stored into a reclosable plastic bag and stored in the incubator (Fig. 1b).

The storage period was 7 days, but storage in silica gel was still performed after 14 days (in this article, named long-term). The protocol for this clinical trial was approved by the clinical trial ethics review committee of Chiyoda Paramedical Care Clinic (publicly registered at UMIN-CTR, trial number: UMIN000028459). Written informed consent was received from all participants. Ten participants were recruited for the collection of fecal samples based on the following criterion: (1) between 0 and 100 years old. (2) Subjects who don’t regularly use specific medicines. (3) Subjects who show understanding of the study procedures and agreement with participating the study by written informed consent prior to the study. (4) Subjects who does not have current medical history of severe disease. (5) Subjects who have been determined eligible by principal investigator. The characteristics of the subjects were as follows: sex: 6 males, 4 females; and age: 31.3 ± 4.5 years.

### 2.2 DNA and metabolite extraction from fecal samples

DNA and metabolite extraction from fecal samples was performed as follows. The fecal samples in control and room temp. conditions were initially lyophilized using a VD-800R lyophilizer (TAITEC, Saitama, Japan) for at least 16 h. Next, fecal samples were shaken vigorously with 3 mm zirconia beads using a Shake Master (1,500 rpm, 10 min; Biomedical Science Co., Ltd., Tokyo, Japan).

Samples were then suspended in DNA extraction buffer containing 400 μL of a 1% (weight/volume) SDS/TE (10 mM Tris-HCl, 1 mM EDTA; pH 8.0) solution and 400 μL of phenol/chloroform/isoamyl alcohol (25:24:1; NACALAI TESQUE, INC., Kyoto, Japan). The fecal samples in the buffer were further shaken with 0.1 mm zirconia/silica beads using a Shake Master (1,500 rpm, 5 min). After centrifugation (17,800 × g; 5 min; room temperature) of the samples, bacterial genomic DNA was purified by the standard phenol/chloroform/isoamyl alcohol protocol.

RNA was removed from the sample by RNase A (Takara Bio, Inc., Shiga, Japan) treatment, and then DNA samples were purified again by the standard phenol/chloroform/isoamyl alcohol protocol. The extracted DNA was shotgun sequenced by Macrogen Japan. After generating the library using a TruSeq Nano, sequencing was performed using HiSeqX (Illumina, San Diego, CA, USA) according to the manufacturer’s protocol. For the 16S rRNA gene analysis, the primers 27F-mod (AGRGTTTGATYMTGGCTCAG) and 338R (TGCTGCCTCCCGTAGGAGT) were used to amplify the V1-V2 region of the 16S rRNA gene [17]. The amplified DNA was sequenced using a MiSeq (Illumina, San Diego, CA, USA) according to the manufacturer’s protocol. The 16S rRNA gene was sequenced at Bioengineering Lab. Co., Ltd. (Kanagawa, Japan). Metabolite extraction from fecal samples was performed based on previously described methods [18]. After extraction of metabolites, the concentration of metabolites was measured by capillary electrophoresis time-of-flight mass spectrometry (CE-TOFMS). The peaks from the CE-TOFMS were identified, and the relative peak areas, which are values based on comparison with the internal standard peak areas, were calculated. From the relative peak area, the quantitative values of some of the metabolites were calculated by comparison with the reference material.

### 2.3 Bioinformatics and statistical analysis

The 16S rRNA gene data were analyzed by QIIME2 (version 2019.10) with our analytical pipeline [19], as follows. Quality filtering and denoising were used to generate amplicon sequence variants (ASVs) by DADA2 (options: --p-trim-left-f 20 --p-trim-left-r 19 --p-trunc-len-f 240 --p-trunc-len-r 140). The ASVs were assigned to taxa by applying the Silva SSU Ref Nr 99 (version 132) classifier (command: “qiime feature-classifier classify-sklearn”; options: default).

Metagenomic shotgun data were processed with the bioBakery3 workflow (version 3.0.0.a.7) to generate functional gene profiles (options: wmgx --bypass-strain-profiling) [20]. The workflow starts with low-quality reads or human genome read removal with KneadData version 0.10.0. Functional gene and metabolic pathway profiles were generated with HUMAnN version 3.0.0.alpha.3. The ChocoPhlAn3 and UniRef 90 databases (obtained January 2019) were used for functional gene annotation, while metabolic pathway annotation was performed with MetaCyc. Functional gene and metabolic pathway profiles were normalized according to relative abundance with humann_renorm_tables.

For statistical analysis, in-house Python scripts were used (version 3.7.6). For the comparison of the distance of the fecal microbiome and metabolome profiles, the Friedman test and Nemenyi test were used (scipy 1.5.2 and scikit_posthocts 0.7.0). In the subsequent analysis, microbes with a mean relative abundance below 0.001 and microbial pathways and metabolites undetected in 75% of the samples were excluded. For the pairwise comparison of the abundances of microbes, microbial pathways, and metabolites, the Wilcoxon signed-rank test with Benjamini−Hochberg false discovery rate (FDR-BH) correction was used (scipy version 1.5.2 and statsmodels 0.10.0). Spearman’s correlation coefficient and the test for no correlation were used to compare the ranks of each microbe, microbial pathway, and metabolite. The relative abundances of microbiome, microbial pathway table, and metabolome abundance table are shown in Supplementary Tables S1-3, respectively.

## 3 Results

### 3.1 Qualitative analysis of the fecal microbiome and metabolome with each sampling method

In this study, we hypothesized that fecal drying with silica gel may preserve the fecal microbiome and metabolome and compared stool drying with silica gel and several other fecal storage methods (Fig. 1). However, metabolome analysis after processing with RNAlater could not be performed because of the high salt content. In addition, the FTA card could not measure each metabolite amount but measure the concentration due to the low stool volume.

First, qualitative analysis was performed. From all samples, 241 genera, 458 metabolic pathways, were detected and 316 metabolites were detected. According to a comparison of the bacteria and metabolites detected by each sampling method, the overlap with the control method was calculated (Table 1). Although microbes and microbial pathways were reasonably well preserved by all methods (microbes: 71.9%-87.2%, microbial pathways: 90.0%-94.3%), metabolites were not well preserved with FTA card storage (36.6 ± 5.3%). Of all sampling methods, silica gel had the highest overlap with the control for genus (86.2 ± 3.1%), microbial pathways (94.3 ± 3.0%) and metabolites (81.3 ± 5.4%). Long-term silca gel preservation retains more fecal features for genus (87.2 ± 3.1%) and metabolites (82.4 ± 4.9%).

**Table 1.**
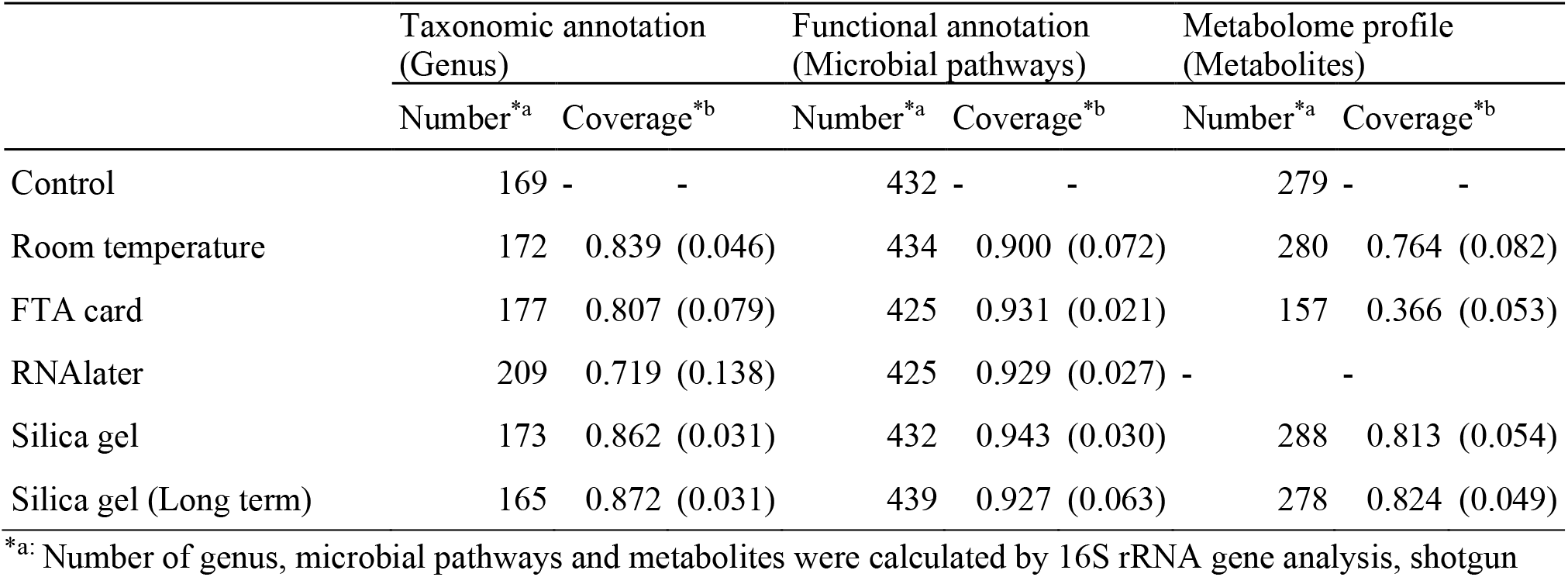
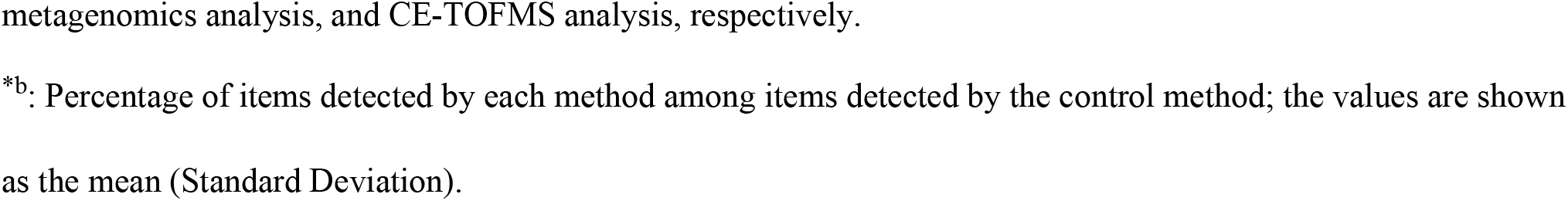
Qualitative analysis of bacteria and metabolites for each sampling method

|  | Taxonomic annotation<br>(Genus) |  | Functional annotation<br>(Microbial pathways) |  | Metabolome profile<br>(Metabolites) |  |
| --- | --- | --- | --- | --- | --- | --- |
|  | Number<sup>*a</sup> | Coverage<sup>*b</sup> | Number<sup>*a</sup> | Coverage<sup>*b</sup> | Number<sup>*a</sup> | Coverage<sup>*b</sup> |
| Control | 169 | - | 432 | - | 279 | - |
| Room temperature | 172 | 0.839 (0.046) | 434 | 0.900 (0.072) | 280 | 0.764 (0.082) |
| FTA card | 177 | 0.807 (0.079) | 425 | 0.931 (0.021) | 157 | 0.366 (0.053) |
| RNAlater | 209 | 0.719 (0.138) | 425 | 0.929 (0.027) | - | - |
| Silica gel | 173 | 0.862 (0.031) | 432 | 0.943 (0.030) | 288 | 0.813 (0.054) |
| Silica gel (Long term) | 165 | 0.872 (0.031) | 439 | 0.927 (0.063) | 278 | 0.824 (0.049) |

<sup>\*a</sup>: Number of genus, microbial pathways and metabolites were calculated by 16S rRNA gene analysis, shotgun
metagenomics analysis, and CE-TOFMS analysis, respectively.
<sup>\*b</sup>: Percentage of items detected by each method among items detected by the control method; the values are shown as the mean (Standard Deviation).

### 3.2 Quantitative analysis

#### 3.2.1 Beta diversity analysis - Precision and accuracy

In measurement, precision (how close measurements are to each other) and accuracy (how close measurements are to the true value) should be high. In this section, we analyzed the precision and accuracy of the determination of the overall microbiome, microbial pathways, and metabolome profile.

First, each method’s precision for microbiome and metabolome profiling was determined. The precision of the microbial pathway profiling was not calculated because it was a singular. The precision of the microbiome profiling was similar for all groups (Fig. 2A). However, the precision of the metabolome profiling was significantly lower in the FTA group than in the other groups (Fig. 2B; *p* < 0.05; Nemenyi test). These results suggest that the microbiome and metabolome profiles were calculated precisely by many methods, except for the metabolome profile when the FTA card method was used.

**Fig. 2.**
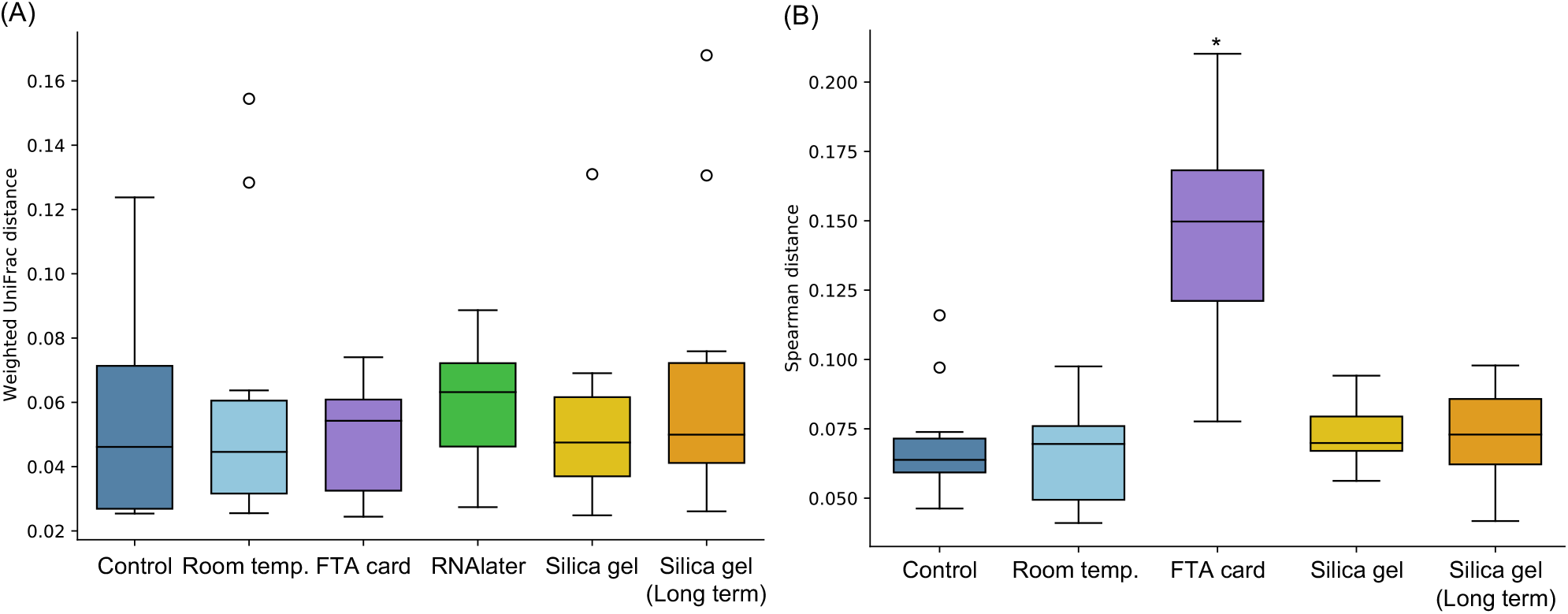
Boxplot of microbiome and metabolome profiling precision. The mean values of the (A) microbiome weighted UniFrac distances, and (B) metabolome spearman distances between triplicates in each condition and subject were calculated and are shown in the boxplots. Microbiome weighted UniFrac distance did not show the significant difference by multiple comparison procedure (Friedman test). *, significant difference from other conditions by Nemenyi test (*p* < 0.05). Room temp., room temperature.

Subsequently, the accuracy of each method was determined (Fig. 3). In determining accuracy, the control method (immediate processing) was treated as the true value. As the positive control, average distances between control samples was used, but there is no positive control for microbial pathways abundance table as it use single sample. As the result, significant differences in microbiome and metabolome profile were detected in Room temp, FTA card, and RNAlater method, excluding RNAlater method in metabolome profile, when compared to controls (Fig. 3A, C).

**Fig. 3.**
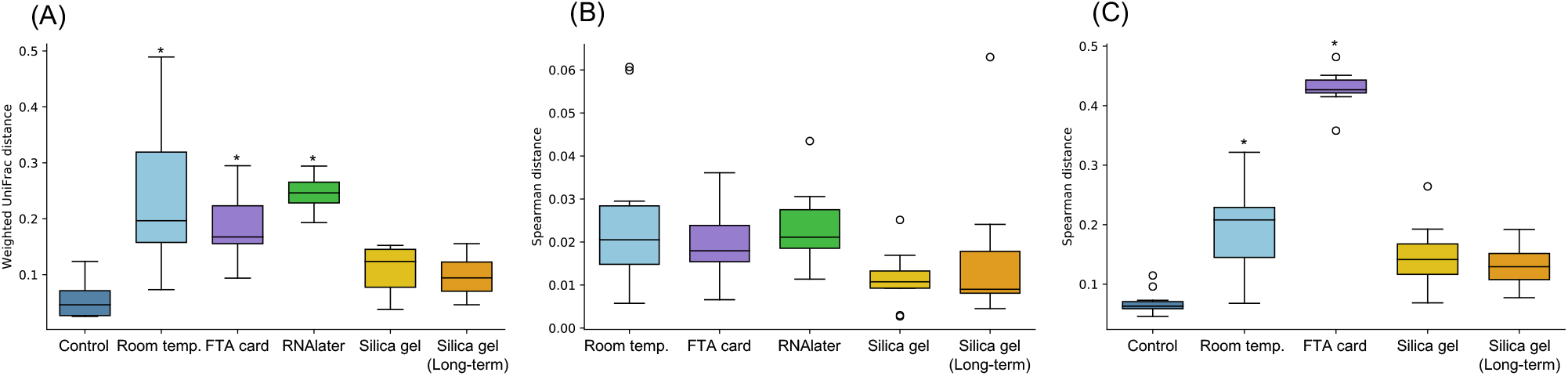
Boxplot of the accuracy of the microbiome, microbial pathway, and metabolome profiling shows that the silica gel preservation method preserves the fecal microbiome and metabolome, resulting in high-accuracy determination of the profiles. The (A) microbiome weighted UniFrac distance, (B) microbial pathway Spearman distance, and (C) metabolome Spearman distance to the control condition in each condition were calculated; for the control condition, the mean value of the distance between replicates was calculated. *, *p* < 0.05 compared with the control in the Nemenyi test.

#### 3.2.2 Differences in the abundance of individual microbes, microbial pathways, and metabolites due to storage methods

Next, abundance on each microbe, microbial pathway, and metabolite was compared to evaluate the effects of storage methods. Although the differences in microbes and microbial pathways were not large, metabolites were depleted in the FTA card method (Fig. 4). For each item, statistical tests were performed between the control condition and each condition, and significant differences were detected for many of the items, but the differences were not significant after FDR adjustment. However, for the RNAlater method, some significant differences were found in microbial pathways abundance even after FDR adjustment; for the FTA card method, several significant differences were found in genus, microbial pathways, and metabolites even after FDR adjustment (Supplementary Tables 4 and 5).

**Fig. 4.**
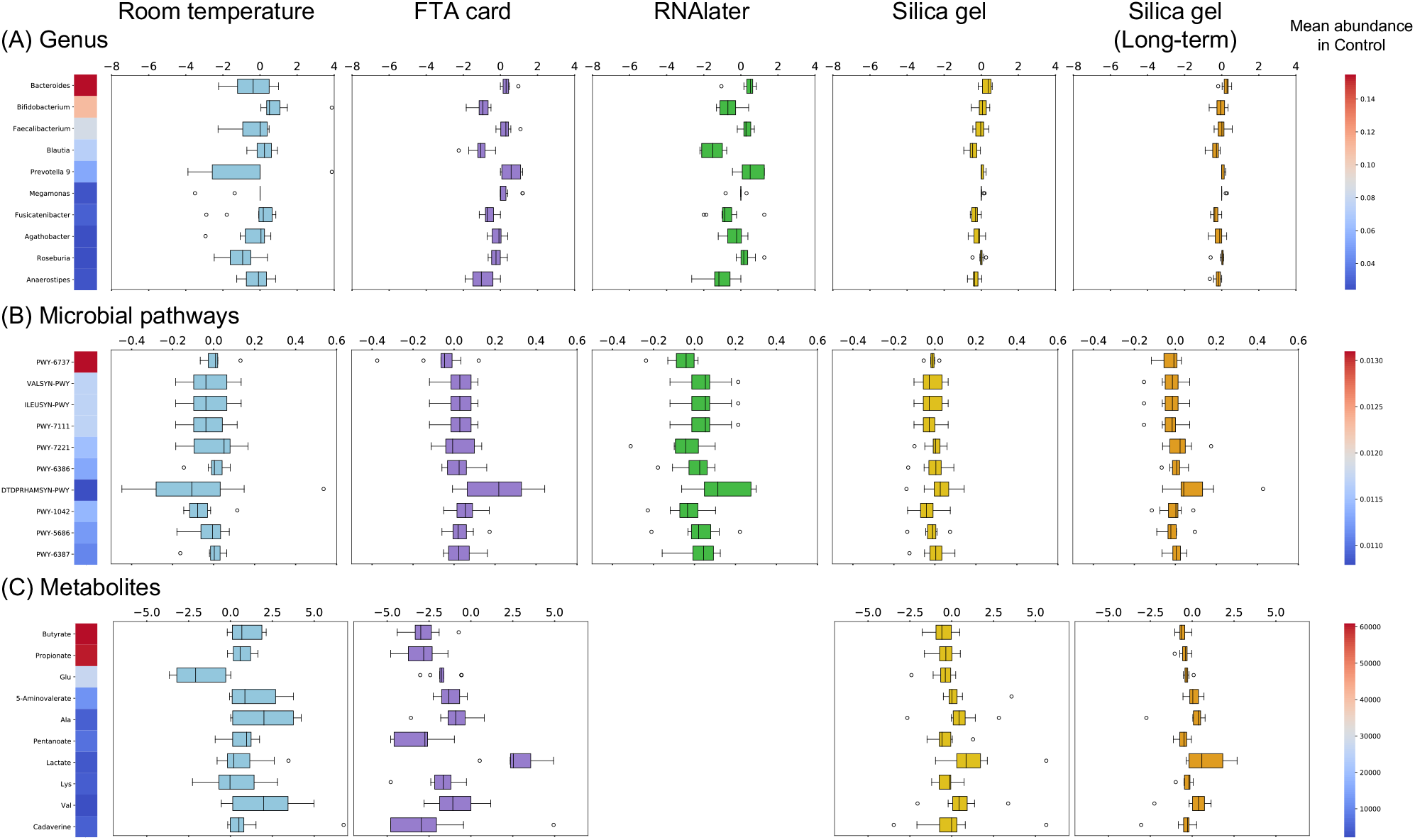
Distribution of the fold change of the abundance of each microbe, microbial pathway, and metabolite compared to that in the control condition. A boxplot of the fold change of the abundance of each microbe, microbial pathway, and metabolite compared to that in the control condition is shown. The 10 items with the highest mean value in all samples were used. For the FTA card method, the fold change was calculated after conversion to relative abundance, as different units were used.

#### 3.2.3 Differences in the rank of individual microbes, microbial pathways, and metabolites due to storage methods

In the previous section, we analyzed the abundances of each microbe, microbial pathway, and metabolite. If there was no difference in abundance, the results with one method could be directly compared to results with a control method. However, if the within-sampling method rankings were consistent across conditions, the same results would be expected in a case−control study within each condition. For each item, we calculated the correlation between the within-control method rankings and the rankings within each method (Fig. 5 and Supplementary Tables 6 and 7). Overall, genus showed a high correlation with control method for all methods,however, fewer metabolites showed significant correlations compared to genus (genus: 92.4%-100%, metabolites: 9.5%-76.2%; Supplementary Tables 6 and 7).

**Fig. 5.**
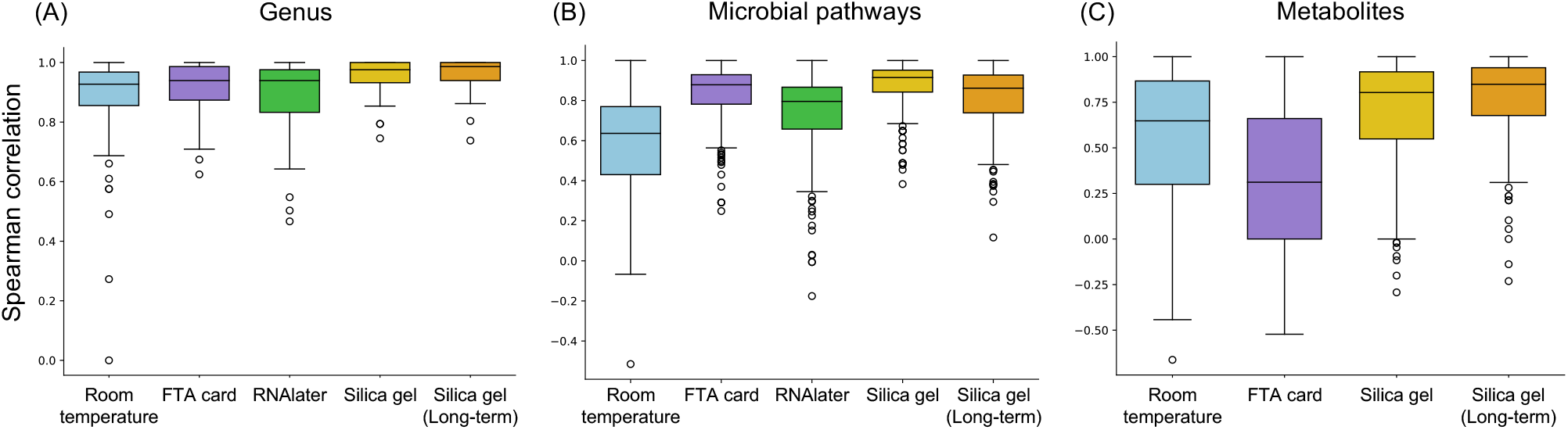
The silica gel method preservation preserves the intra-ranking of genus, microbial pathway, and metabolite abundance. The Spearman correlation coefficients between the control method and each method for each genus, microbial pathway, and metabolite are shown in the boxplots.

A comparison of the results with the different methods showed that the highest number of bacteria and metabolites were preserved when fecal samples were stored in silica gel (Fig. 5a-c). In addition, the room temperature condition was characterized by a low correlation coefficient for *Bacteroides* (Spearman’s Rho = 0.273; Supplementary Table 6). The Spearman correlation coefficients, *p*-values, and q-values for each microbe, microbial pathway and metabolite are listed in Supplementary Table 6.

## 4 Discussion

The purpose of this study was to explore methods of preserving microbes, microbial genes and metabolites in fecal samples. Among various methods, we focused particularly on fecal sample storage by drying. To evaluate the microbes, microbial genes, and metabolites in fecal samples, we performed a comparative analysis with several methods.

First, fecal samples that were preserved with RNAlater could not measure metabolome contents by CE-TOFMS due to high salt content. In addition, it is standard to normalize by dry/wet stool weight when measuring metabolite concentrations [21]. However, the FTA card could not be used to measure stool weight due to the low stool volume. These methods are not considered well suited for measuring metabolome abundance.

Next, we analyzed (1) qualitative detection, (2) quantitative precision, and (3) quantitative accuracy for the microbiome, metagenome, and metabolome profiling in fecal samples. In the (1) qualitative analysis, there were no large differences among all the methods conducted in this study for the preservation of microbes and microbial pathways. However, there were differences regarding metabolites; the FTA card method resulted in fewer metabolites detected (36.6 ± 5.3% compared to the control method). It is known that there are local and microenvironmental differences in the microbiome depending on the location of the stool [22]. Although stool samples were mixed as a pretreatment in this study, it is possible that the metabolites included in the FTA card were biased due to the small amount of stool in the FTA card. In the (2) quantitative precision analysis, it was shown that the microbiome and microbial pathways were preserved with all methods, but the metabolome had low precision with the FTA card method. The same reason noted above may regarding large technical replication. In the (3) quantitative accuracy analysis, the difference between the technical replicates of the control method was smaller than the difference between the control method and each method in terms of median (Fig. 3). In some methods except for silica gel method, significant differences were detected. This result suggests that each sampling method affects the result of microbiome, metagenome, and metabolome profiles.

For the analysis of each microbe, microbial pathway, and metabolite, (1) comparative quantity analysis and (2) comparative rank analysis for each method were performed. The comparative quantity analysis indicate that the values are comparable even if the methods are different. The comparative rank analysis indicates whether the results obtained from comparisons within the same method are the same. In the comparison analysis of the quantities, significant differences were detected for some genus, microbial pathways, and metabolites with all methods. In particular, the FTA card method detected significant differences in the abundances of some items even after FDR correction. (2) Many genera, microbial pathways, and metabolites were found to be conserved in the rank analysis. This result suggested that comparisons such as case−control studies within the same method would yield the same results. However, fewer metabolites showed significant correlations compared to genus. When discussing the results of metabolite analysis that are not significantly correlated, we should be aware of the possibility that differences may appear depending on the method (Supplementary Tables 6 and 7). Focusing on individual items, the rankings of individual microbe and microbial pathway abundance were relatively conserved. However, for the room temperature method, the low correlation of *Bacteroides* (Spearman’s Rho = 0.273; Supplementary Table 6) may have influenced the relative abundance of other bacteria.

Therefore, the room temperature method may not be suitable for gut microbiome analysis. In addition, only 76.2% of metabolites were conserved in the abundance of ranking, even with the best method (Long-term silica gel preservation; Fig. 5c and Supplementary table 7). Especially focusing on propionate and butyrate, two of the most commonly measured SCFA metabolites, they both strongly correlated with the silica gel method (butyrate, room temperature: 0.285, FTA card: 0.539, silica gel: 0.830, silica gel (long-term): 0.952; propionate, room temperature: 0.697, FTA card: 0.340, silica gel: 0.830, silica gel (long-term): 0.952; Supplementary Figure 1).

The following limitations should be considered for our study. First, we analyzed multivariate data, which included abundances of hundreds of intestinal microbes, microbial pathways, and metabolites. Therefore, there is a multiple comparison test problem. It is thus necessary to examine each microbe, microbial pathway, and metabolite in another larger cohort. Second, only healthy subjects were used in this study. Although the silica gel drying method will be applied to research on bacteria specific to sick subjects in the future, we did not analyze the gut microbiome and metabolome of sick subjects. Further validation of the gut microbiome and metabolome of diseased individuals may be necessary.

In conclusion, fecal sample drying could be used to preserve the fecal microbiome and metabolome. This drying method could make a significant contribution to microbiome research when used in larger cohorts.

## Data Availability

The obtained sequence data are available in the DNA Data Bank of Japan (DDBJ) Sequence Read Archive (DRA) (DRA accession number: DRA016258).

## 5 Conflict of Interest

T.N., Y.Y., Y.N., Y.T., M.I., F.S., K.F., and S.M. are employees of Metagen, Inc.; T.Y. and S.F. are founders of Metagen, Inc. Metagen, Inc. sells commercially a fecal sampling kit using silica gel as MGKit.

## 6 Author Contributions

Conceptualization: T.N. and S.F.; Formal Analysis: Y.Y., Y.N., F.S., and K.F.; Investigation: Y.T., M.I. and T.N.; Visualization: Y.Y. and Y.N.; Writing – Original Draft: Y.N.; Writing – Review & Editing: T.N., Y.Y., Y.N., Y.T., M.I., S.M., T.Y., and S.F.

## 7 Funding

This study was funded by Metagen, Inc.

## 8 Acknowledgments

The supercomputing resource was provided by the Human Genome Center (University of Tokyo). Additionally, we would like to thank all the members who discussed the study and participated in the trial at Metagen, Inc.

## 11 Supplementary Figure

**Supplementary Figure 1.**
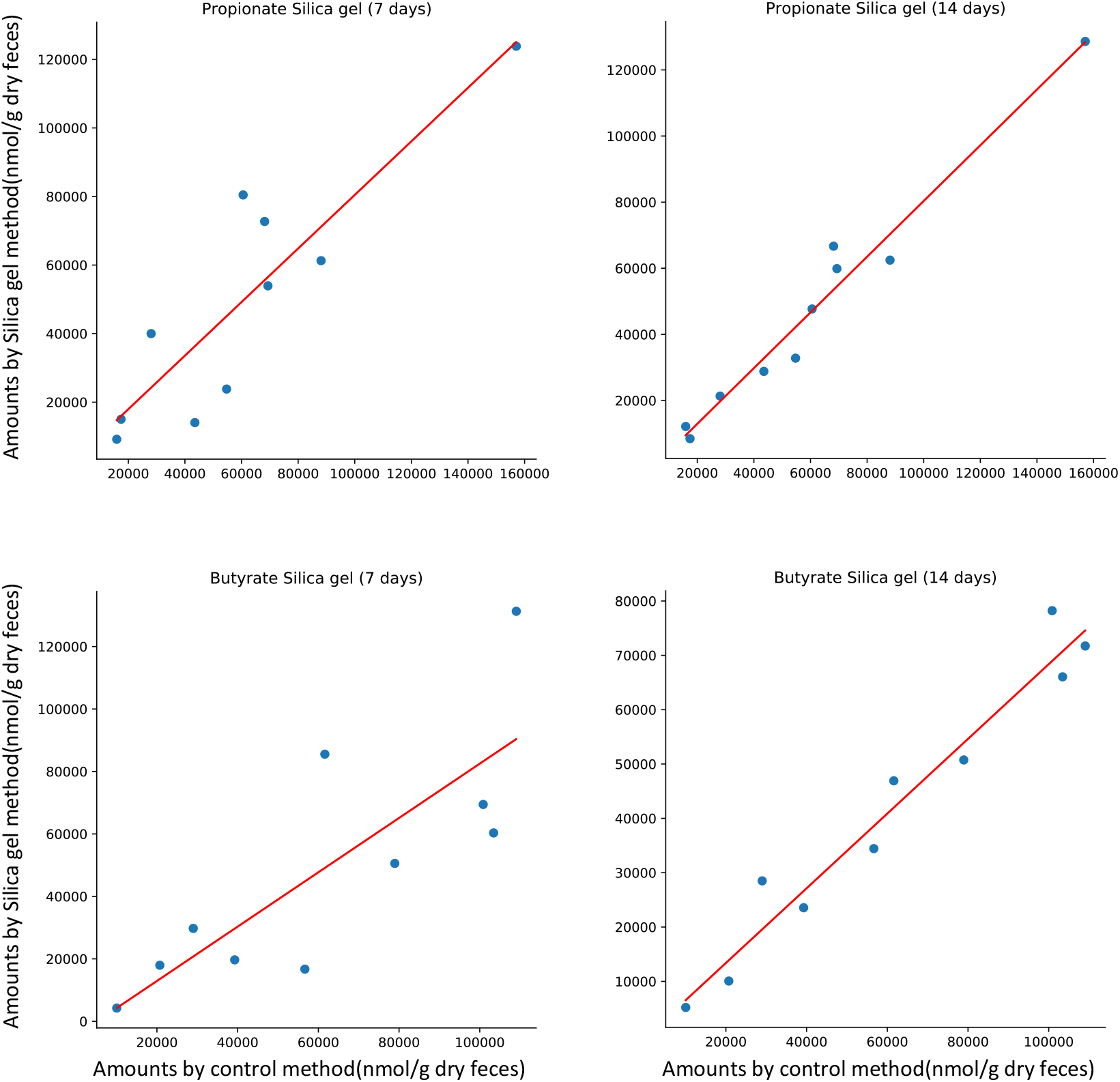
Scatter plot of propionate and butyrate in control and silica gel method. Scatter plots were drawn using the mean triplicate values for each subject. The red line represents the regression curve.

## Notes

### Competing Interest Statement

The authors have declared no competing interest.

### Funding Statement

This study did not receive any funding

### Author Declarations

IRB of Chiyoda Paramedical Care Clinic gave ethical approval for this work

## References

1. Lloyd-Price J, Arze C, Ananthakrishnan AN, Schirmer M, Avila-Pacheco J, et al. Multi-omics of the gut microbial ecosystem in inflammatory bowel diseases. Nature. 2019;569:655–62.

2. Dawkins JJ, Allegretti JR, Gibson TE, McClure E, Delaney M, Bry L, et al. Gut metabolites predict Clostridioides difficile recurrence. Microbiome. 2022;10:87.

3. Wirbel J, Pyl PT, Kartal E, Zych K, Kashani A, Milanese A, et al. Meta-analysis of fecal metagenomes reveals global microbial signatures that are specific for colorectal cancer. Nat Med. 2019;25:679–89.

4. Koh A, Molinaro A, Ståhlman M, Khan MT, Schmidt C, Mannerås-Holm L, et al. Microbially Produced Imidazole Propionate Impairs Insulin Signaling through mTORC1. Cell. 2018;175:947-961.e17.

5. Tang WHW, Wang Z, Levison BS, Koeth RA, Britt EB, Fu X, et al. Intestinal Microbial Metabolism of Phosphatidylcholine and Cardiovascular Risk. N Engl J Med. 2013;368:1575–84.

6. Jalanka J, Salonen A, Salojärvi J, Ritari J, Immonen O, Marciani L, et al. Effects of bowel cleansing on the intestinal microbiota. Gut. 2015;64:1562–8.

7. Nishimoto Y, Mizutani S, Nakajima T, Hosoda F, Watanabe H, Saito Y, et al. High stability of faecal microbiome composition in guanidine thiocyanate solution at room temperature and robustness during colonoscopy. Gut. 2016;65:1574–5.

8. Liang Y, Dong T, Chen M, He L, Wang T, Liu X, et al. Systematic Analysis of Impact of Sampling Regions and Storage Methods on Fecal Gut Microbiome and Metabolome Profiles. mSphere. American Society for Microbiology; 2020;5:e00763–19.

9. Guan H, Pu Y, Liu C, Lou T, Tan S, Kong M, et al. Comparison of Fecal Collection Methods on Variation in Gut Metagenomics and Untargeted Metabolomics. mSphere. American Society for Microbiology; 2021;6:e00636–21.

10. Chirife J, del Pilar Buera M, Labuza TP. Water activity, water glass dynamics, and the control of microbiological growth in foods. Critical Reviews in Food Science and Nutrition. Taylor & Francis; 1996;36:465–513.

11. ⟨1112⟩ Application of Water Activity Determination to Nonsterile Pharmaceutical Products [Internet]. Available from: https://doi.usp.org/USPNF/USPNF_M402_01_01.html

12. Chase MW, Hills HH. Silica gel: An ideal material for field preservation of leaf samples for DNA studies. TAXON. 1991;40:215–20.

13. Allison MJ, Round JM, Bergman LC, Mirabzadeh A, Allen H, Weir A, et al. The effect of silica desiccation under different storage conditions on filter-immobilized environmental DNA. BMC Research Notes. 2021;14:106.

14. Kimura I, Ozawa K, Inoue D, Imamura T, Kimura K, Maeda T, et al. The gut microbiota suppresses insulin-mediated fat accumulation via the short-chain fatty acid receptor GPR43. Nat Commun. 2013;4:1829.

15. Hee B van der, Wells JM. Microbial Regulation of Host Physiology by Short-chain Fatty Acids. Trends in Microbiology. Elsevier; 2021;29:700–12.

16. Ueyama J, Oda M, Hirayama M, Sugitate K, Sakui N, Hamada R, et al. Freeze-drying enables homogeneous and stable sample preparation for determination of fecal short-chain fatty acids. Analytical Biochemistry. 2020;589:113508.

17. Kim S-W, Suda W, Kim S, Oshima K, Fukuda S, Ohno H, et al. Robustness of Gut Microbiota of Healthy Adults in Response to Probiotic Intervention Revealed by High-Throughput Pyrosequencing. DNA Research. 2013;20:241–53.

18. Kim Y-G, Sakamoto K, Seo S-U, Pickard JM, Gillilland MG, Pudlo NA, et al. Neonatal acquisition of Clostridia species protects against colonization by bacterial pathogens. Science. 2017;356:315–9.

19. Bolyen E, Rideout JR, Dillon MR, Bokulich NA, Abnet CC, Al-Ghalith GA, et al. Reproducible, interactive, scalable and extensible microbiome data science using QIIME 2. Nat Biotechnol. 2019;37:852–7.

20. Beghini F, McIver LJ, Blanco-Míguez A, Dubois L, Asnicar F, Maharjan S, et al. Integrating taxonomic, functional, and strain-level profiling of diverse microbial communities with bioBakery 3. Turnbaugh P, Franco E, Brown CT, editors. eLife. eLife Sciences Publications, Ltd; 2021;10:e65088.

21. So D, Whelan K, Rossi M, Morrison M, Holtmann G, Kelly JT, et al. Dietary fiber intervention on gut microbiota composition in healthy adults: a systematic review and meta-analysis. Am J Clin Nutr. 2018;107:965–83.

22. Hosomi K, Ohno H, Murakami H, Natsume-Kitatani Y, Tanisawa K, Hirata S, et al. Method for preparing DNA from feces in guanidine thiocyanate solution affects 16S rRNA-based profiling of human microbiota diversity. Sci Rep. 2017;7:4339.

